# Expanding the role of village malaria workers in Cambodia: implementation and evaluation of four health education packages

**DOI:** 10.1101/2023.03.10.23287110

**Authors:** Mipharny Betrian, Dafne Umans, Moul Vanna, Sam Ol, Bipin Adhikari, Chan Davoeung, James J Callery, Yok Sovann, Thomas J Peto, Richard J Maude, Rob W van der Pluijm, Voeunrung Bunreth, Martin P. Grobusch, Michèle van Vugt, Yoel Lubell, Lorenz von Seidlein, Arjen M Dondorp, Siv Sovannaroth, Dysoley Lek, Rupam Tripura

## Abstract

**Background:** Early access to correct diagnosis and appropriate treatment is essential for malaria elimination, and in Cambodia this relies on village malaria workers (VMWs). Decreasing malaria transmission leave VMWs with diminished roles. Activities related to the control of other health conditions could keep these community health workers relevant.

**Methods:** During 2022, 120 VMWs attended training at local health centres on four health education packages: 1. hygiene and sanitation; 2. disease surveillance; 3. management of mild illness; 4. vaccination and antenatal care. All training and evaluation sessions were documented through meeting minutes, and 19 focus group discussions (FGDs) were conducted among VMWs and health centre personnel. Audio-records of FGDs were transcribed and translated in English and underwent thematic analysis.

**Results:** VMWs reported strong interest in the training and welcomed the expansion of their roles thus assuring their continued relevance. VMWs prioritized disease surveillance and management of mild illness among the available training packages because these topics were seen as most relevant. While training was considered comprehensible and important, the low literacy among VMWs was an impediment suggesting training materials need to be delivered visually. Since VMWs have limited resources, incentives could ensure that VMWs are motivated to undertake additional roles and responsibilities.

**Conclusions:** The transformation of VMWs into community health workers with roles beyond malaria is a promising approach for sustaining health care provision in remote areas. Training needs to consider the low scientific literacy, time constraints and limited resources of VMWs.

## Introduction

The Greater Mekong Subregion (GMS) in Southeast Asia had historically a high burden of febrile illnesses, including malaria [1]. However, since the millennium there has been a substantial decrease in malaria transmission in the region. To prevent the further evolution and spread of antimalarial drug resistance in the GMS, concerted efforts are underway to eliminate all human malaria by 2030 [2, 3]. Early, accurate diagnosis and effective treatment is the current main strategy for malaria elimination. In the GMS, as in most of Southeast Asia, the success of this strategy depends on networks of community health workers (CHWs) [4].

Due to a shortage of healthcare professionals in rural areas, local volunteers provide basic health services in many low or lower middle-income countries (LMICs). These CHWs networks can improve access to testing and treatment despite a lack of sophisticated medical training [5]. In some places CHWs provide a broad range of health services [6]. In Cambodia CHWs work in vertical programmes, such as for malaria control. As malaria transmission diminishes in Southeast Asia, the importance of other febrile illnesses caused by non-malarial pathogens increases. Since village malaria workers (VMWs) currently do not manage non-malarial causes of fever, fewer patients attend them, thus limiting their usefulness and relevance. Identifying new duties, diagnostics, and activities to evolve VMW networks into CHW networks will extend their spectrum of usefulness. Expansion of the roles of CHWs beyond malaria has been applied successfully elsewhere and resulted in an increased number of patients tested for malaria, and in strengthened rural healthcare services [6].

In Cambodia, VMWs are tasked with health promotion, diagnosis and treatment of malaria [7, 8]. Currently, VMWs are unable to manage patients once malaria has been ruled out [4]. There are multiple healthcare providers in Cambodian villages including government health care centres, non-governmental organizations (NGOs) that provide health information, and village health support groups (VHSG) who educate community members and brief healthcare personnel on prevalent health conditions in the villages. If VMW are to expand their roles, they must be integrated into these existing structures. To meet Cambodian malaria elimination targets, it is essential that febrile patients continue to attend VMWs until local elimination is achieved and the risk of re-importation subsides. We report here a research project which provided short health education courses on non-malaria activities to Cambodian VMWs, and a qualitative assessment of the acceptability and feasibility of this approach for sustaining and strengthening the VMW programme.

## Methods

### Study context

This study is part of operational research supported by the Global Fund to Fight AIDS, Tuberculosis and Malaria entitled *Sustaining village health worker programmes with expanded roles in the GMS*. In Cambodia, this included a project to explore the feasibility of expanded roles of VMWs, of which this study was a part. It was a collaboration between Mahidol Oxford Tropical Medicine Research Unit (MORU) and Action for Health Development Cambodia (AHEAD) and was supervised by the Cambodian National Center for Entomology, Parasitology and Malaria Control (CNM). AHEAD led the field implementation of the study and this study reports activities from June to December 2022. During the study, VMWs were offered training on various diseases in four districts of Battambang province. The training and its evaluation were conducted primarily using qualitative methods.

### Participants

VMWs and health centre personnel were chosen from four districts (Samlout, Koas Krala, Pailin and Rukh Kiri) in Battambang province. Nine health centres were included where most of the training and evaluation were conducted. The VMWs were selected for the current training regardless of their age, experience, or socio-demographic background. Personnel employed at the nine health centres were invited to provide feedback regarding the health education training. Written informed consents from all potential participants were obtained before the interview/discussion sessions.

### Training packages

Four broad topics for the health education packages were established in advance by AHEAD in consultation with CNM and partners. Based on the discussions with health centre personnel and VMWs, the topics were refined to reflect the prevalences of diseases within these rural areas (excluding malaria). Four innovative health education packages were developed, covering the following topics: 1. hygiene and sanitation; 2. disease surveillance (subtopics: COVID-19, dengue); 3. management of mild illness (subtopics: typhoid fever, respiratory infection, first aid and dehydration); 4. vaccination and antenatal care (ANC). The training sessions for each topic were conducted at local health centres and each session had a duration of 90-120 minutes. Training was given in small groups led by a trainer from AHEAD with occasional support from health centre chiefs, and the content was based on a lesson guide. Communication was mostly verbal with some use of posters, flipcharts, and materials from previous health education activities. These training materials contained key messages in text and pictures.

Training was structured into one of the topics (and subtopics) and were delivered primarily orally with examples of the conditions, consequences and potential ways to manage them. Training sessions were also interactive and included the opportunity for questions and participation by attendees, for example role play during first aid training.

### Data collection and analysis

A total of 14 FGDs were conducted at the nine health centres during the implementation phase of the study (June to August 2022) and these FGDs took place immediately after health education workshops. This was done to capture the initial impression from the VMWs of the health education. During the evaluation phase (from October 2022), five FGDs with VMWs, organised one to two months after they had received training on the topic. Additionally, one FGD was conducted with community members.

Each FGD had a duration of 30 to 60 minutes. The FGDs were conducted in Khmer and facilitated by two members of the AHEAD supervisory team, with six to eleven participants per session. All audio recordings were translated and transcribed into English and were cross-checked by the local interviewers and investigators. No information was collected that could identify individual participants during or after data collection.

All audio transcripts were transcribed and translated to English, and then collated into qualitative data analysis software: NVivo version 12 by QSR international, Australia, and into Microsoft Excel. The analyses were conducted using a deductive method that used an existing codebook derived from the interview guide to inform the data coding. Further codes that emerged from reading the transcripts line-by-line were added into the existing codebook. At all stages, data analysis was conducted under the supervision of a social scientist (BA). Any disagreements on coding process, codes and corresponding descriptions were resolved by discussions and, at times, by seeking input from interviewers. Major themes and sub-themes from the coded data and corresponding data are the basis of the results presented below.

## Results

### Demographic characteristics of participants

The study population was 102 (91.1%) VMWs and 10 (8.9%) health centre personnel. The majority of participants (105; 93.8%) were more than 30 years old. 57.1% (n=57) of the participants were females. The highest education level of the participants was primary level (25.9%), secondary level (34.8%) and high school (30.4%) for the VMWs. The remaining 8.9% of participants had a higher-level education corresponding to their specialization such as laboratory technician, midwife, or nurse. During the evaluation phase, four FGDs were held with 41 (98%) VMWs and 1 (2%) health centre personnel. The majority of the participants were female (n=24; 57%) and more than 30 years old (n=38, 90%). **[Table 1]**

**Table 1.** Sociodemographic Characteristics of Participants.

|  |  | Implementation<br>phase | Evaluation<br>phase |
| --- | --- | --- | --- |
| **Gender (n,%)** |  |  |  |
|  | Female | 64 (57.1%) | 24 (57%) |
|  | Male | 48 (42.9%) | 18 (43%) |
| **Age (n,%)** |  |  |  |
|  | ≤ 30 | 7 (6.2%) | 4 (10%) |
|  | > 30 | 105 (93.8%) | 38 (90%) |
| **Education level (n,%)** |  |  |  |
|  | Primary school | 29 (25.9%) | 27 (64%) |
|  | Secondary School | 39 (34.8%) | 7 (17%) |
|  | High School | 34 (30.4%) | 8 (19%) |
|  | Other | 10 (8.9%) | - |
| **Occupation (n,%)** |  |  |  |
|  | Healthcare staff | 10 (8.9%) | 1 (2%) |
|  | VMW | 102 (91.1%) | 41 (98%) |
| **Distribution per health centre (n,%)** |  |  |  |
|  | Kra Chab | 17 (15.2%) | 8 (19%) |
|  | O'Chra | 6 (5.5%) | 3 (7%) |
|  | Soun Koma | 4 (3.6%) | 3 (7%) |
|  | Prey Tralach | 8 (7.1%) | - |
|  | Koas Kralor | 28 (25%) | 10 (24%) |
|  | Boeung Run | 14 (12.5%) | 11 (26%) |
|  | Chark Roka | 7 (6.3%) | - |
|  | Kampong Lpov | 10 (8.9%) | 7 (17%) |
|  | Tasahn | 18 (16.1%) | - |

### Expanding the roles of VMWs

The majority of the VMWs and health centre personnel were satisfied with the proposed expansion of the roles of the VMWs, which seemed to serve their motivation to continue improving their level of skills and contribute to their community. Although adding responsibilities to their existing role would increase their workload and could be burdensome, the VMWs felt that their sense of communal responsibilities outweighed the workload.

> *I: When we add more tasks or roles to the village malaria workers, do they have more workload?*
>
> *R: Frankly, there are quite a lot of tasks for us. It is a big workload. It is a burden, but we need to do it for the health of the community*
>
> *-* FGD with sixteen VMWs and one health centre worker, Koas Kralor

Adding roles and responsibilities to VMWs and their services to the community were further justified because community members reported VMWs as the preferred source of health care. VMWs were consulted first and patients had an opportunity to discuss and decide whether to visit health centre or a referral hospital. Their preference to consult VMWs were also rooted in their proximity, good relationship and familiarity.

> *I: Why did they decide to go to the VMW as the first service provider?*
>
> *R: Because he lives so close to us, so we feel warm. On the other hand, he frequently visits every house in the village when he does not go to work*.
>
> -FGD with eight community members, Prey Tralach

### Training packages

According to the VMWs and health centre staff the health information from the training sessions were clear and covered the most important points, namely cause, symptoms, treatment, and prevention for the diseases.

> *I: Do you think what we have learned is useful or not and if it is, how is it useful?*
>
> *R: It is good because we help explain to them and also help let them understand the manifestations, the symptoms, the care and the prevention*
>
> - FGD with six VMWs and one health centre worker, Chark Roka

Nonetheless, there were suggestions to improve the training. Participants suggested to include longer, and more comprehensive training in future. The VMWs also requested receiving more feedback from facilitators throughout the training sessions.

> *I: Do you think the content is acceptable, with each topic shown with a few pictures and the time?*
>
> R: *We would like the opportunity for a two-day training course and to practice providing education for each other, this way we are prepared when we conduct it in the community*
>
> *-* FGD with twelve VMWs and two health centre workers, Boeung Run

The majority requested more resources, such as pamphlets and leaflets to provide education to the community members. More visual materials were requested, such as images or posters because some VMWs have only limited literacy.

> *I: Regarding the methodology you received from the trainer to use to disseminate to people in the community, how do you improve on it?*
>
> *R: We need to have enough teaching materials to show the community. If we do not have materials, they will not understand what we teach them*
>
> - FGD with twenty-six VMWs and two health centre workers, Koas Kralor

During the evaluation, VMWs reiterated the importance of visual aids in training. VMWs suggested the need for physical materials such as pictures, posters or leaflets presentations that could be referred to if and when required. Training on visualizing the health topics, including formal coursebooks, were also recommended. VMWs recommended broadcast media such as radio or television, and megaphones for wider information sharing.

### Recommended topics for training

The most common preferences for future education topics amongst the participants were frequently encountered non-communicable diseases, such as diabetes, hypertension, arthritis and gout.

> *I: I want you to clarify in terms of teaching people in the community, which diseases do you want to focus on? The diseases which you think may occur in the future or currently, but no one notices it?*
>
> *R: High blood pressure and diabetes, we do not know how to prevent them and would like to receive knowledge to instruct the population on how to prevent these*
>
> - FGD with twelve VMWs and two health centre workers, Boeung Run

VMWs showed interest in additional tasks such as measuring blood pressure and glucose levels. Health centre personnel thought that measuring blood pressure was a straightforward task that VMWs could complete, suggesting VMWs could potentially serve as a link between the health centres and community by monitoring these diseases.

> *I: What other activities or topics would you like to learn, or know, besides what we have learned or described just now?*
>
> *R: I would like to request a blood pressure monitor and heartbeat monitor, since there are plenty of elderly people in the villages with high blood pressure, who frequently ask me if I can measure their blood pressure*
>
> - FGD with twelve VMWs and two health centre workers, Boeung Run

During the evaluation phase as well, VMWs echoed some of the recommendations they shared at the outset. In response to how they would like to see their roles in the future and what topics they preferred, a mix of both communicable (e.g. tuberculosis, cholera, and the common cold) and non-communicable (e.g. diabetes and hypertension) diseases was recommended. Some VMWs shared increased workload could be a concern when expanding their roles. It was reiterated that financial support is critical to compensate for the time invested. Another challenge was a possible overlap of new roles allocated to VMWs with the pre-existing roles of other health volunteers.

### Training implementation in the community

VMWs disseminated health education to their communities in several ways. Larger gatherings of people were found to be ineffective because VMWs found communicating in big groups difficult. The majority of respondents preferred training in small groups consisting of three to four participants. VMWs conducted health education when the village chief would gather villagers for a meeting or after ceremonies and funerals at the pagoda, and some preferred to educate community members individually, for example.

> *I: The mode of education? You conduct education differently, right?*
>
> *R: It is easier with a small group of people, for example one or two people, when we see people, we just start talking and provide information. In most cases, when we conduct education, it is after people visit us for a test, just two to three people face to face. If education is conducted in a large group, they do not value us*.
>
> - FGD with six VMWs and one health centre worker, Chark Roka
>
> *I: What about during the holiday seasons such as New Year’s Day or Buddha Day? Can you conduct dissemination then?R: Every Sunday elderly people gather to do exercise and they also have a meeting every month. I always take the opportunity to teach them during these gatherings. I also do it at funerals or on Buddha Day*
>
> - FGD with twenty-six VMWs and two health centre workers, Koas Kralor

### Challenges

The VMWs reported their incentives to be too low to carry out their roles and responsibilities, specifically when they had to travel to engage community members, which prevented them from carrying out other tasks. Incentives also enhanced motivation of community members to participate. For instance, community members were less interested in engaging in health education sessions if there was no incentive, such as snacks, soap, gifts or other materials related to the theme of the education session.

> *I: What about during the holiday seasons such as New Year’s Day or Buddha Day? Can you conduct dissemination then?*
>
> *R: It is quite impossible to gather community members and teach them unless there is something to give them as an incentive*.
>
> - FGD with twenty-six VMWs and two health centre workers, Koas Kralor
>
> *I: Well, is there anything else?*
>
> *R: If there is a budget, then it would be a little better but if it is too scarce then it does not even cover the gasoline fee back and forth*
>
> - FGD with eight VMWs and two health centre workers, Kampong Lpov

It was difficult to retain the participants’ attention and presence when providing health education. Some participants did not take the health education sessions seriously. It was challenging to assemble participants because of their competing priorities, particularly farmers during the peak of farming season. Most of the participants in meetings organized by VMWs were female, and absence of men was attributed to farm work.

> *I: When you conducted awareness raising, did you ever have issues with participants, for example did they follow your advice, or consider it important?*
>
> *R: Some of them did not follow our advice, mostly because they thought we did not have enough knowledge to educate them*
>
> - FGD with seventeen VMWs and one health centre worker, Tasanh
>
> *I: When do you think it is an effective time to conduct dissemination regarding health education?*
>
> *R: During this season people often go to the mountains to harvest crops, so we rarely see them because they have already gone to the plantation. Since it is harvesting season, they have all gone to the plantation and it is hard to gather them for a meeting*.
>
> - FGD with twenty-six VMWs and two health centre workers, Koas Kralor

### Reflections by VMWs on training packages

VMWs felt that the training packages had potential to prevent disease in the community, and were pleased because these topics were chosen by them and the community members. Training packages were felt to enhance community members’ ability to deal with certain symptoms and diseases without needing to visit health workers. VMWs reported that additional support from other respected members of the community or health centre staff would be useful to circulate the information they learned during the education sessions to the wider community. Village leaders, monks, health centre staff and teachers were most often named as potential aides to strengthen their message and training to community members. Nonetheless, VMWs expressed that health education should be conducted with their involvement, as they would otherwise be seen as ‘unnecessary’ and potentially lose status and trust from community members.

The VMWs kept records of their dissemination activities within their communities and these were reviewed by research staff. Self-reported data from the VMWs’ logbooks showed a total of 6,192 community members (59% female) had received training on health topics from the VMWs. There were differences in the numbers of community members who were reached during subsequent dissemination of the health topics by VMWs. Disease surveillance was the most commonly discussed topic whereas vaccination & ANC was the topic on which the least amount of community members (n=481) were educated [**table 2**]. A total of 6,001 community members were educated on one of the four topics included in the training.

**Table 2.** Health education dissemination to community members by village malaria workers.

| Topic | Sub-topics | Participants |
| --- | --- | --- |
| Hygiene & Sanitation |  | 1315 |
| Disease Surveillance |  | 2803 |
|  | Dengue | 1834 |
|  | COVID-19 | 969 |
| Disease Management |  | 1402 |
|  | Typhoid fever | 485 |
|  | First-aid | 43 |
|  | Respiratory infection | 365 |
|  | Dehydration | 509 |
| EPI & ANC |  | 481 |
|  | EPI | 280 |
|  | ANC | 201 |

## Discussion

This study reports practicalities associated with the implementation and evaluation of training on health topics among VMWs and community members in rural western Cambodia. The training packages were found to be relevant because of their potential to expand VMWs’ roles. Simple, visual, and physical training materials are needed for the training to communicate with an audience with limited literacy.

VMWs networks have been an important resource to implement training packages and were found to yield positive outcomes in Myanmar, Vietnam and Bangladesh [9-11].

The majority of the VMWs and health centre personnel were positive about the expansion of the roles of the VMWs. Several VMWs acknowledged that adding responsibilities to their existing role would increase their workload because of the competing responsibilities they had, however, they also saw the health-related responsibilities more as a service to the community. Adding new roles and responsibilities was thought to promote their relevance and hence their sense of recognition by the community [12, 13]. Indeed, in recent mass drug administration and malaria chemoprophylaxis activities in Southeast Asia, CHWs have played an important role [14, 15]. In Cambodia, like in many LMICs, CHWs are the first point of contact for the majority of health problems [12, 16]. There is growing international evidence of the potential benefits of seeking health care from CHWs in terms of cost, coverage, quality of services, and reductions in disease morbidity and mortality [17-21]. CHWs and VMWs have an inherent interest to be of value, thus want to expand their roles and responsibilities [12, 13, 16].

Despite the basic nature of the content, it was difficult for the participants to understand and retain all the key points contained in the health information they received. Many VMWs recommended providing them with physical training materials such as flow charts, pamphlets, or leaflets, so that they could retain and reflect on the content. Elsewhere in rural Southeast Asia low literacy and language barriers have been highlighted as challenges to health communication [22, 23]. Visualization has been proven to increase attention, recall, comprehension and adherence of audiences with low literacy levels [24]. This limitation in literacy highlights the need to enhance the clarity and visualization of the information presented [22, 23]. Even among those who are fully literate, many participants only have primary or secondary school education, and some concepts of basic biology and disease transmission are not easy to communicate to them [25].

VMWs reported some of the challenges of the training packages that included brief training, lack of refresher training; and recommended longer and frequent training sessions which have been demonstrated to be beneficial by the previous studies [4, 26-28]. One-off training may be inadequate and could be counterproductive. A need for repeated training was also highlighted by a recent study among VMWs in Kravanh district, Cambodia [12]. Refresher messages (training) have also been an important strategy to strengthen understanding and retention among the audience [29].

VMWs also reported challenges in carrying out training in their community, specifically the lack of incentives to compensate for their time was a major concern. In this study, the incentives were inadequate to complete all the training activities. VMWs used their own resources such as vehicles and gasoline. Inadequate compensation can be a major problem and has been reported in other countries e.g. Bhutan and Uganda [28, 30, 31]. Both financial and non-financial incentives have been shown to promote productivity by CHWs [30-32].

To reinforce VMWs’ credibility when offering health education to community members, local health centre staff or local authority figures could be mobilized to garner respect from community members. ‘Authority engagement’ has been shown to be an important element of achieving community participation [22, 33]. Apart from engaging with the authorities and community leaders, engaging and liaising with health workers is vital as they bear ‘institutional’ and ‘inter-personal’ trust among the community members because they depend on them for health-related matters [34-36].

VMWs and community members recommended other health topics that they would like to be trained on in future. Such recommendations were motivated by their own desire to be relevant for wider diseases in the community and community members’ motivation to expand their knowledge base. These recommendations suggest that VMWs are open to new health topics and demonstrate their readiness to undertake new responsibilities. Nonetheless, studies caution about balancing the responsibilities given to VMWs, particularly to not compromise the quality of services by overburdening them [37, 38]. Striking a balance between disease coverage and quality of service therefore requires further operational studies.

### Strengths and limitations

To make up for delays during COVID-19 restrictions, the initial implementation of training happened quickly, therefore local health centre staff were not trained in advance to lead the education sessions and local NGO staff had to deliver the training. Due to time constraints, visual health education materials were not available for all topics, which could have enhanced the sessions. Language barriers between international members of the research team, local staff, and study participants, limited the communication during FGDs.

Based on stakeholder feedback, future education topics to be considered for further expansion of VMWs roles could focus on non-communicable diseases, as these are prevalent in most populations and could be useful in preventing long lasting complications. Multiple projects elsewhere have successfully equipped volunteers with the necessary tools and education to conduct monitoring of non-communicable diseases [11, 39-41]. Promising tasks might include measuring blood pressure, measuring blood glucose levels and assisting community members with taking or administering medication.

## Conclusion

This study explored the implementation and feedback of health training packages among village malaria workers in Cambodia. Training on health topics can successfully be provided to this population with the utilization of visual aids, and flowcharts with simplified information and tasks. Training on health topics was well-received by the participants and their dissemination to their communities represents a promising and feasible new role for them. The study findings imply that training sessions should be interactive and offered repeatedly, and fair incentives should be provided for VMWs to take on new responsibilities.

## Data Availability

Data collected for the study, including deidentified individual participant data and a data dictionary defining each field in the set, will be made available to others. These data will be available with publication via the MORU data sharing committee, Chairperson, Professor Phaikyeong Cheah. Access will be provided for analysis by bona fide researchers with or without investigator support, after approval of a proposal, and upon a signed data access agreement.

## Declarations

### Ethical approval

Ethics approval was obtained from the National Ethics Committee for Health Research Cambodia (NECHR 0125), the Oxford Tropical Research Ethics Committee (OxTREC ref 517-21), and the study is registered on clinicaltrials.gov (NCT05045547).

## Acknowledgements

We thank the study participants, and the staff who conducted the study at health centres in Pailin and Battambang provinces, with special thanks to Lim Vanthy and Bou Sakun who facilitated the focus group discussions, and to Sazid Zaman, Jacklyn Adella and Marc Visser.

## Author contributions

MB, DU, TJP, BA, RT and JC conceived the study. MB, DU, MV, SO, BA, RT, JC and TJP led the implementation and data collection. MB, DU, and BA conducted the data analysis followed by discussion with JC, RT and TJP. MB, DU, BA, and TJP wrote the first draft of the manuscript. All authors read, revised, and approved the final version of the manuscript.

---

**Figure 1.**
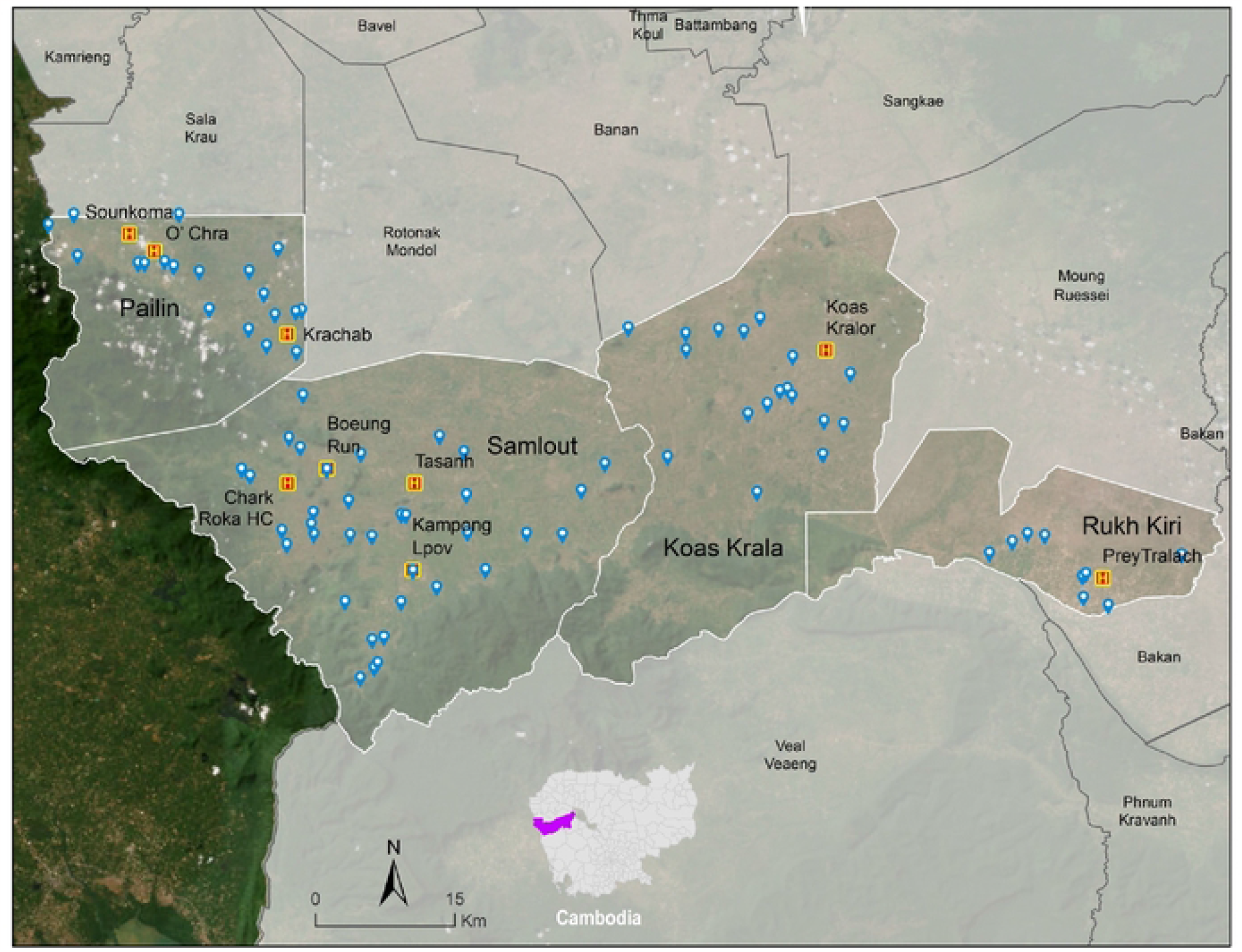
Location map of operational districts, health centres and villages included in the study. Blue pins represent villages where VMWs reside. Yellow ‘H’ symbols represent health centres where training and focus group discussions took place.

**Figure 2.**
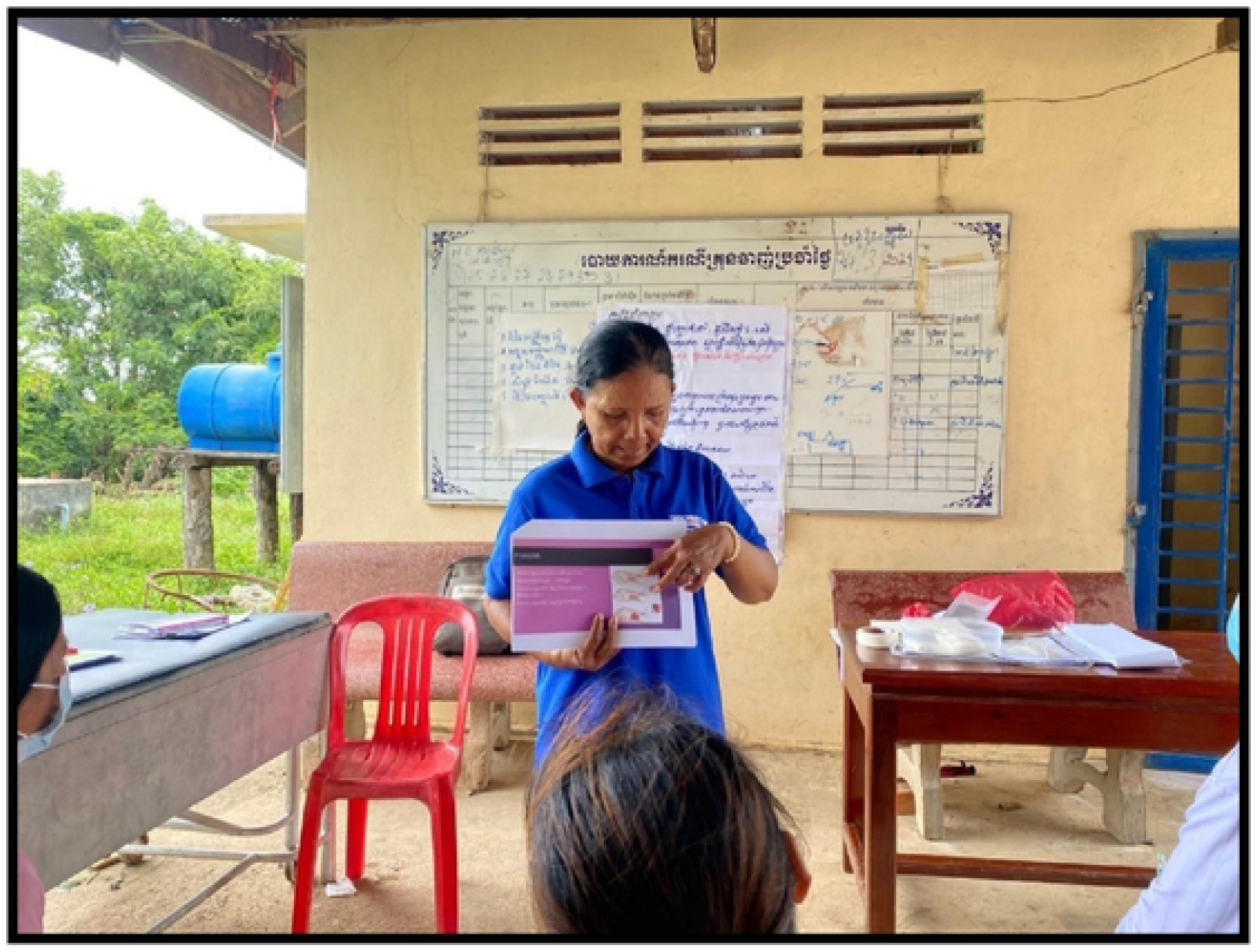
Health education training.

**Figure 3.**
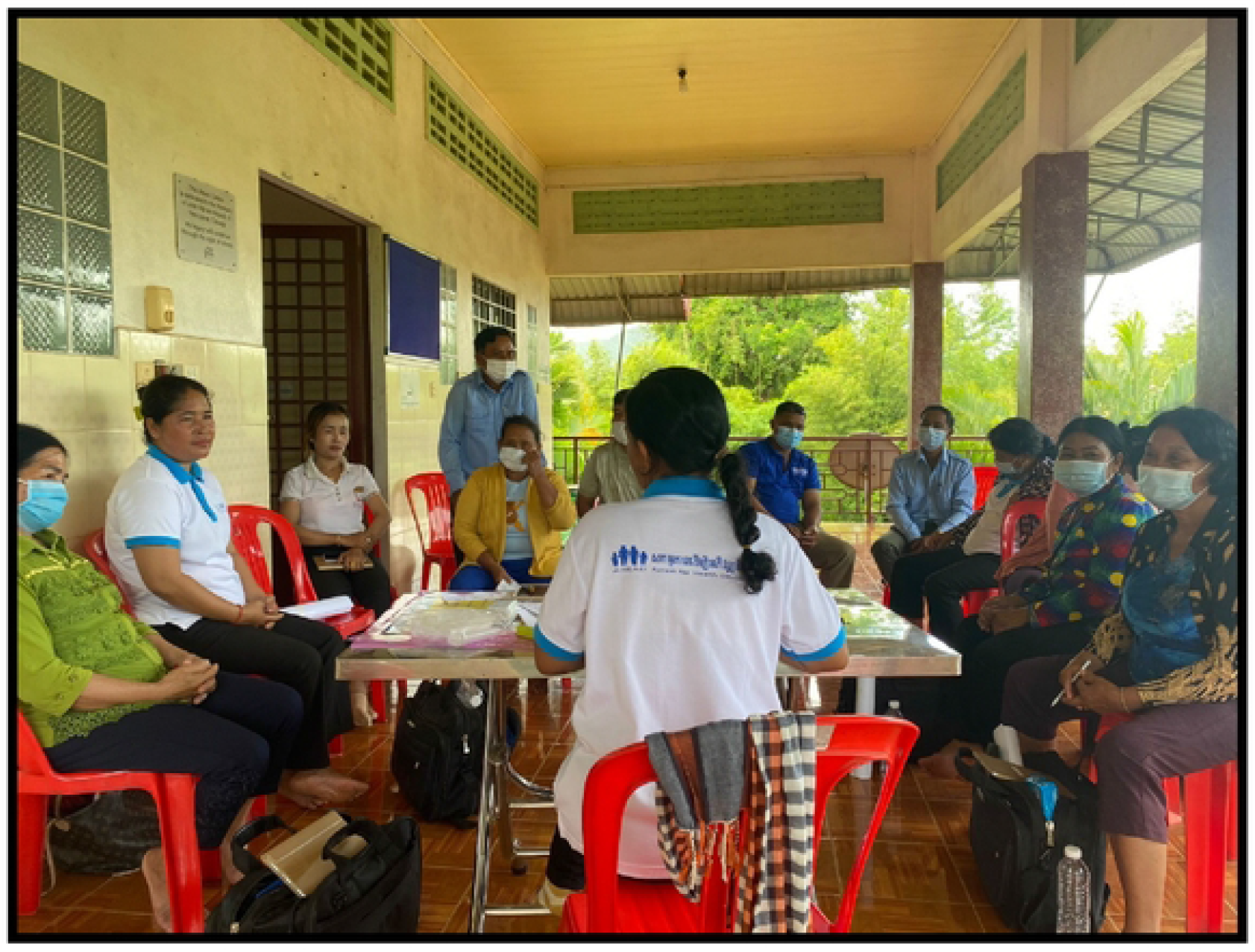
Focus Group Discussion.

## References

1. Chhim S, Piola P, Housen T, Herbreteau V, Tol B: Malaria in Cambodia: A Retrospective Analysis of a Changing Epidemiology 2006–2019. Int J Environ Res Public Health 2021, 18.

2. Organization WH: Strategy for malaria elimination in the Greater Mekong Subregion: 2015–2030. Manila: WHO Regional Office for the Western Pacific; 2015.

3. Organization WH: World malaria report 2022. World Health Organization; 2022.

4. Canavati SE, Lawpoolsri S, Quintero CE, Nguon C, Ly P, Pukrittayakamee S, Sintasath D, Singhasivanon P, Peeters Grietens K, Whittaker MA: Village malaria worker performance key to the elimination of artemisinin-resistant malaria: a Western Cambodia health system assessment. Malar J 2016, 15:282.

5. Benskin LL: A concept development of the village health worker. Nurs Forum 2012, 47:173–182.

6. McLean ARD, Wai HP, Thu AM, Khant ZS, Indrasuta C, Ashley EA, Kyaw TT, Day NPJ, Dondorp A, White NJ, Smithuis FM: Malaria elimination in remote communities requires integration of malaria control activities into general health care: an observational study and interrupted time series analysis in Myanmar. BMC Med 2018, 16:183.

7. Ashley EA, Dhorda M, Fairhurst RM, Amaratunga C, Lim P, Suon S, Sreng S, Anderson JM, Mao S, Sam B, et al: Spread of artemisinin resistance in Plasmodium falciparum malaria. N Engl J Med 2014, 371:411–423.

8. National Center for Parasitology EaMCC, Cambodia: Cambodia Malaria Elimination Action Framework 2021–2025. Available online at https://www.cnm.gov.kh/userfiles/Cambodia%20Malaria%20Elimination_English%20FINAL.pdf (Accessed on 21st March, 2022). 2020.

9. Win Han O, Hoban E, Gold L, Kyu Kyu T, Thazin L, Aung T, Fowkes FJI: Optimizing Myanmar’s community-delivered malaria volunteer model: a qualitative study of stakeholders’ perspectives. Malar J 2021, 20:79.

10. Long H, Ma Z, Hanh TTD, Minh HV, Rawal LB, Urmi DS, Jafar TH, Tang S, Abdullah AS: Engaging village health workers in non-communicable disease (NCD) prevention and control in Vietnam: A qualitative study. Glob Public Health 2020, 15:611–625.

11. Rawal L, Jubayer S, Choudhury SR, Islam SMS, Abdullah AS: Community health workers for non-communicable diseases prevention and control in Bangladesh: a qualitative study. Glob Health Res Policy 2020, 6:1.

12. Adhikari B, Tripura R, Dysoley L, Callery JJ, Peto TJ, Heng C, Vanda T, Simvieng O, Cassidy-Seyoum S, Ley B, et al: Glucose 6 Phosphate Dehydrogenase (G6PD) quantitation using biosensors at the point of first contact: a mixed method study in Cambodia. Malar J 2022, 21:282.

13. Kok MC, Broerse JEW, Theobald S, Ormel H, Dieleman M, Taegtmeyer M: Performance of community health workers: situating their intermediary position within complex adaptive health systems. Hum Resour Health 2017, 15:59.

14. Tripura R, von Seidlein L, Sovannaroth S, Peto TJ, Callery JJ, Sokha M, Ean M, Heng C, Conradis-Jansen F, Madmanee W, et al: Antimalarial chemoprophylaxis for forest goers in southeast Asia: an open-label, individually randomised controlled trial. Lancet Infect Dis 2022.

15. von Seidlein L, Peto TJ, Landier J, Nguyen TN, Tripura R, Phommasone K, Pongvongsa T, Lwin KM, Keereecharoen L, Kajeechiwa L, et al: The impact of targeted malaria elimination with mass drug administrations on falciparum malaria in Southeast Asia: A cluster randomised trial. PLoS Med 2019, 16:e1002745.

16. Adhikari B, Tripura R, Peto TJ, Callery JJ, von Seidlein L, Dysoley L, Dondorp AM: Village malaria workers for the community-based management of vivax malaria. The Lancet Regional Health-Southeast Asia 2023, 9:100128.

17. Yeboah-Antwi K, Pilingana P, Macleod WB, Semrau K, Siazeele K, Kalesha P, Hamainza B, Seidenberg P, Mazimba A, Sabin L, et al: Community case management of fever due to malaria and pneumonia in children under five in Zambia: a cluster randomized controlled trial. PLoS Med 2010, 7:e1000340.

18. Kalyango JN, Rutebemberwa E, Karamagi C, Mworozi E, Ssali S, Alfven T, Peterson S: High adherence to antimalarials and antibiotics under integrated community case management of illness in children less than five years in eastern Uganda. PLoS One 2013, 8:e60481.

19. Christopher JB, Le May A, Lewin S, Ross DA: Thirty years after Alma-Ata: a systematic review of the impact of community health workers delivering curative interventions against malaria, pneumonia and diarrhoea on child mortality and morbidity in sub-Saharan Africa. Hum Resour Health 2011, 9:27.

20. McCord GC, Liu A, Singh P: Deployment of community health workers across rural sub-Saharan Africa: financial considerations and operational assumptions. Bull World Health Organ 2013, 91:244–253b.

21. Vaughan K, Kok MC, Witter S, Dieleman M: Costs and cost-effectiveness of community health workers: evidence from a literature review. Hum Resour Health 2015, 13:71.

22. Adhikari B, Pell C, Phommasone K, Soundala X, Kommarasy P, Pongvongsa T, Henriques G, Day NPJ, Mayxay M, Cheah PY: Elements of effective community engagement: lessons from a targeted malaria elimination study in Lao PDR (Laos). Glob Health Action 2017, 10:1366136.

23. Kajeechiwa L, Thwin MM, Nosten S, Tun SW, Parker D, von Seidlein L, Tangseefa D, Nosten F, Cheah PY: Community engagement for the rapid elimination of malaria: the case of Kayin State, Myanmar. Wellcome Open Res 2017, 2:59.

24. Houts PS, Doak CC, Doak LG, Loscalzo MJ: The role of pictures in improving health communication: a review of research on attention, comprehension, recall, and adherence. Patient Educ Couns 2006, 61:173–190.

25. Priest S: Critical science literacy: What citizens and journalists need to know to make sense of science. Bulletin of Science, Technology & Society 2013, 33:138–145.

26. Zheng C, Anthonypillai J, Musominali S, Chaw GF, Paccione G: Community Perceptions of Village Health Workers in Kisoro, Uganda. Ann Glob Health 2021, 87:82.

27. O’Brien MJ, Squires AP, Bixby RA, Larson SC: Role development of community health workers: an examination of selection and training processes in the intervention literature. Am J Prev Med 2009, 37:S262–269.

28. Musinguzi LK, Turinawe EB, Rwemisisi JT, de Vries DH, Mafigiri DK, Muhangi D, de Groot M, Katamba A, Pool R: Linking communities to formal health care providers through village health teams in rural Uganda: lessons from linking social capital. Hum Resour Health 2017, 15:4.

29. Kahneman D: Thinking, fast and slow. Macmillan; 2011.

30. Tshering D, Tejativaddhana P, Siripornpibul T, Cruickshank M, Briggs D: Identifying and confirming demotivating factors for village health workers in rural communities of Bhutan. Int J Health Plann Manage 2018, 33:1189–1201.

31. Tshering D, Tejativaddhana P, Siripornpibul T, Cruickshank M, Briggs D: Motivational Factors Influencing Retention of Village Health Workers in Rural Communities of Bhutan. Asia Pac J Public Health 2019, 31:433–442.

32. Zheng CY, Musominali S, Chaw GF, Paccione G: A Performance-Based Incentives System for Village Health Workers in Kisoro, Uganda. Ann Glob Health 2019, 85.

33. Peto TJ, Tripura R, Davoeung C, Nguon C, Nou S, Heng C, Kunthea P, Adhikari B, Lim R, James N, et al: Reflections on a community engagement strategy for mass antimalarial drug administration in Cambodia. Am J Trop Med Hyg 2018, 98:100–104.

34. Adhikari B, Yeong Cheah P, von Seidlein L: Trust is the common denominator for COVID-19 vaccine acceptance: A literature review. Vaccine X 2022, 12:100213.

35. Gilson L: Trust and the development of health care as a social institution. Soc Sci Med 2003, 56:1453–1468.

36. Molyneux CS, Peshu N, Marsh K: Trust and informed consent: insights from community members on the Kenyan coast. Soc Sci Med 2005, 61:1463–1473.

37. Kruk ME, Gage AD, Arsenault C, Jordan K, Leslie HH, Roder-DeWan S, Adeyi O, Barker P, Daelmans B, Doubova SV, et al: High-quality health systems in the Sustainable Development Goals era: time for a revolution. Lancet Glob Health 2018, 6:e1196–e1252.

38. Jain S: India’s army of unrecognised, unpaid female health workers. BMJ 2021, 375:n2509.

39. Widyasari V, Rahman FF, Lin KH, Wang JY: The Effectiveness of Health Services Delivered by Community Health Workers on Outcomes Related to Non-Communicable Diseases among Elderly People in Rural Areas: A Systematic Review. Iran J Public Health 2021, 50:1088–1096.

40. Le HT, Le TA, Mac TD, Nguyen DN, Vu HN, Truong ATM, Quang Do AT, Bui HTT, Do HTT, Nguyen ATH, et al: Non-communicable diseases prevention in remote areas of Vietnam: Limited roles of health education and community workers. PLoS One 2022, 17:e0273047.

41. Raithatha SJ, Kumar D, Amin AA: Training Village Health Workers in Detection and Monitoring of Noncommunicable Diseases: A Low Cost Option for Rural Areas Facing the Emerging Health Epidemic. Fam Community Health 2017, 40:253–257.

